# Evaluating Short-term Forecast among Different Epidemiological Models under a Bayesian Framework

**DOI:** 10.1101/2020.10.11.20210971

**Authors:** Qiwei Li, Tejasv Bedi, Guanghua Xiao, Yang Xie

**Author notes:** The author contributed equally with the first author to this work.

## Abstract

Forecasting of COVID-19 daily confirmed cases has been one of the several challenges posed on the governments and health sectors on a global scale. To facilitate informed public health decisions, the concerned parties rely on short-term daily projections generated via predictive modeling. We calibrate stochastic variants of growth models and the standard SIR model into one Bayesian framework to evaluate their short-term forecasts. In summary, it was noted that none of the models proved to be golden standards across all the regions in their entirety, while all outperformed ARIMA in a predictive capacity as well as in terms of interpretability.

## 1 Introduction

COVID-19, a respiratory disease caused by the novel coronavirus SARS-CoV-2, has rapidly become an ongoing global pandemic. It has become the leading cause of death in the United States (U.S.) and is still spreading fast in almost every other country. Given the extent of the physical and economic suffering caused by the pandemic, there is an urgent public health need to better predict the spread of COVID-19 both locally and nationwide. Since the emergence of the COVID-19 outbreak, several predictive modeling approaches have been proposed to predict trends of the disease to come up with effective policies and measures to minimize casualties. Broadly speaking, there are five types of approaches to forecasting the number of new cases or the expected total mortality caused by the COVID-19: 1) time-series forecasting such as autoregressive integrated moving average (ARIMA) (see e.g. Elmousalami and Hassanien, 2020; Perone, 2020); 2) growth curve fitting based on the generalized Richards curve (GRC) or its special cases (see e.g. Batista, 2020; COVID et al., 2020; Roosa et al., 2020; Jia et al., 2020; Wu et al., 2020); 3) compartmental modeling based on the susceptible-infectious-removed (SIR) models or its derivations (see e.g. Fanelli and Piazza, 2020; Kucharski et al., 2020; Li et al., 2020; Liu et al., 2020; Pan et al., 2020; Pei and Shaman, 2020; Song et al., 2020; Sun et al., 2020; Wang et al., 2020; Yamana et al., 2020; Yang et al., 2020); 4) agent-based modeling (see e.g. Gomez et al., 2020); 5) artificial intelligence (AI)-inspired modeling (see e.g. Bullock et al., 2020; Hu et al., 2020; Distante et al., 2020; Vaishya et al., 2020).

Each type of approach, either being deterministic or stochastic, has its own strengths. For instance, the ARIMA model combines the regressive process and the moving average, allowing to predict a given time series by considering its own lags and lagged forecast error. Curve fitting approaches (also known as phenomenological modeling) fit a curve to the observed number of cumulative confirmed cases or deaths with a certain error structure (e.g. Gaussian or Poisson), enabling meaningful interpretation through curve parameters while accounting for measurement errors. Compartmental modeling (also known as mechanistic modeling) approaches consider a partition of the population into compartments corresponding to different stages of the disease and characterize the disease transmission dynamics by the flow of individuals through compartments. Agent-based modeling approaches use computer simulations to study the dynamic interactions among the agents (e.g., people in epidemiology) and between an agent and the environment. AI-based modeling approaches usually embeds time series, clustering, and forecasting under one roof, resulting in an exemplary predictive performance. There has been a growing debate amongst researchers over model performance evaluation and finding the best model appropriate for a certain feature (cases, deaths, etc.), a particular regional level (county, state, country, etc.), and more. Fair evaluation and comparison of the output of different forecasting methods have remained an open question (Tabataba et al., 2017) sine models vary in their complexity in terms of the number of variables and parameters that characterize the dynamic states of the system.

Although a comparison of predictive models for infectious diseases has been discussed in the literature, to our best knowledge, no existing work systematically compares their performances, particularly with the same amount of data information. In this paper, we calibrate stochastic variants of six different growth models (i.e. logistic, generalized logistic, Richards, generalized Richards, Bertalanffy, and Gompertz) and the standard SIR model, all of which can be specified by an ordinary differential equation(ODE), into one flexible Bayesian modeling framework. The main reason that why we limit our analysis to these two modeling approaches is that they can not only make both short and long-term forecasts, but also provide useful insights to understand the disease dynamics of COVID-19. The growth models provide an empirical approach without a specific basis on the mechanisms that give rise to the observed patterns in the cumulative infection data, while the compartmental models incorporate key mechanisms involved in the disease transmission dynamics in order to explain patterns in the observed data.

In our Bayesian modeling framework, the bottom-level is a negative binomial model that directly models infection count data and accounts for the over-dispersed observational errors. The top-level is from a choice of growth or compartmental models that characterizes a certain disease transmission dynamic through ODE(s). The Markov chain Monte Carlo (MCMC) algorithm is used to sample from the posterior distribution. The short-term forecasts are made from the resulting MCMC samples. We perform the rolling-origin cross-validation procedure to compare the prediction error of different stochastic models. In terms of regions, we consider the top 20 countries in terms of confirmed case numbers for a country-level analysis. It is observed that 1) as the models learned more and more data, the predictive performance improved in general for all the regions; 2) none of the models proved to be golden standards across all the regions in their entirety, while the ARIMA model underperformed all stochastic models proposed in the paper. Besides, we offer a graphical interface that allows users to interact with the future trends of COVID-19 at different geographic locations in the U.S. based on the real-time COVID-19 data. This daily updated web portal is currently being used to inform local policy-makers and the general public.

The rest of the paper is organized as follows. Section 2 introduces a variety of stochastic epidemiological models under a unified Bayesian framework. The detailed MCMC algorithms, posterior inference, and forecasting procedures are described in Section 3. Section 4 mainly discusses an algorithmic approach to measure and compare the predictive performance of the proposed models and the benchmark ARIMA model. Besides, we analyze the COVID-19 data for the top 20 countries with the most confirmed cases, including the U.S. Finally, we conclude with remarks in Section 5 and provide information about implementations in Section 6.

## 2 The Bayesian Framework

In this section, we present a bi-level Bayesian framework for predicting new daily confirmed cases during a pandemic in a closed population. Section 2.2 introduces the bottom level, which directly models the observed counts while accounting for measurement errors. Section 2.3 and 2.4 describe two choices of the top level, characterizing the epidemic dynamics through growth curve or compartmental trajectories, respectively. Before introducing the main components, we summarize the possibly observable data as follows.

### 2.1 Data notations

Let ***C*** = (*C*_1_, …, *C*_*T*_) be a sequence of cumulative confirmed case numbers observed at *T* successive equally spaced points in time (e.g. day) in a specific region, where each entry *C*_*t*_ ∈ ℕ for *t* = 1, …, *T*. let *C*_0_ be the initial value and ***Ċ*** = (*Ċ*_1_, …, *Ċ*_*T*_) be the lag one difference of ***C***, where *Ċ*_1_ = *C*_1_ − *C*_0_ and each following entry *Ċ*_*t*_ = *C*_*t*_ − *C*_*t*−1_, *t* = 2, …, *T*, i.e. the difference between two adjacent observations. In the analysis and modeling of a series of infectious disease daily report data, the time-series data could also be the cumulative death numbers, recovery case numbers, or their sums, denoted by ***D*** (Death), ***E*** (Recovery), and ***R*** (Removed), and their corresponding new case numbers, denoted by 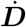, ***Ė***, and 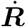. Assuming a closed population with size *N*, the time-series data could also be the number of susceptible people, denoted by ***S***, with each entry *S*_*t*_ = *N* − *C*_*t*_. In reality, only confirmed cases and deaths are reported in most regions. Recovery data are not available or suffer from under-reporting issues even if existing. Thus, our main goal is to make predictions of the future trend of an infectious diseases only based on the confirmed cases ***C***.

### 2.2 Bottom-level: Time-series count generating process

We consider the new case number observed at time *t*, i.e. *Ċ*_*t*_, are sampled from a negative binomial (NB) model,

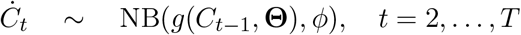

as it automatically accounts for measurement errors and uncertainties associated with the counts. Here, we use NB(*µ, ϕ*), *µ, ϕ* > 0 to denote a NB distribution with expectation *µ* and dispersion 1*/ϕ*. We assume this stochastic process is a Markov process, where the present state (at time *t*) depends only upon its previous state (at time *t* − 1). Therefore, the NB mean is a function, denoted by *g*(*·*), of the case number observed at time *t* − 1, characterized by a set of interpretable/uninterpretable model parameters **Θ**. With this parameterization, the NB variance is *µ* + *µ*^2^*/ϕ*, indicating that *ϕ* controls the variance of measurement error. A small value leads to a large variance to mean ratio, while a large value approaching infinity reduces the NB model to a Poisson model with the same mean and variance. The probability mass function of a NB random variable *Y* is 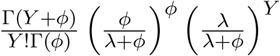. Thus, we can write the full data likelihood as

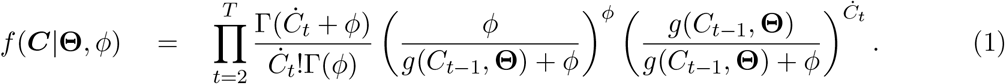

For the prior distribution of the dispersion parameter *ϕ*, we choose a gamma distribution, *ϕ* ∼ Ga(*a*_*ϕ*_, *b*_*ϕ*_). We recommend small values, such as *a*_*ϕ*_ = *b*_*ϕ*_ = 0.001, for a non-informative setting (Gelman et al., 2006). Note that the proposed framework can be also viewed as a stochastic discrete-time state-space model with a negative binomial error structure. The proposed Bayesian models, on average, mimics the epidemic dynamics and is more flexible than those deterministic epidemiological models, as it accounts for measurement error and has the potential to incorporate existing information into the prior structure of **Θ**.

### 2.3 Top-level I: Growth model

We first discuss the choices of *g*(*·*) when implementing growth models. The development of a variety of growth curves originates from population dynamics (Haberman, 1998) and growth of biological systems (Werker and Jaggard, 1997; Desta et al., 1999; Kaps et al., 2000; Topal and Bolukbasi, 2008) modeling. A number of growth curves have been adapted in epidemiology for trend characterization and forecasting of an epidemic, such as the severe acute respiratory syndrome (SARS) (Hsieh et al., 2004; Hsieh, 2009), dengue fever (Hsieh and Ma, 2009; Hsieh and Chen, 2009), pandemic influenza A (H1N1) (Hsieh, 2010), Ebola virus disease (EVD) (Chowell et al., 2014; Pell et al., 2018), Zika fever (Chowell et al., 2016), and COVID-19 (Batista, 2020; Jia et al., 2020; Lee et al., 2020; Wu et al., 2020).

The underlying assumption is that the rate of growth of a population, organism, or infectious individuals eventually declines with size. The logistic curve (also known as sigmoid curve) is the simplest growth curve of continuous time *u* ∈ ℝ. It is a non-negative symmetric ‘S’-shaped curve with equation 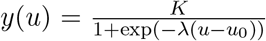, where *u*_0_ is the midpoint, *K* is the maximum value, and *λ* reflects the steepness of the curve. It is clear to see that *y*(*u*) approaches *K* when *u* → ∞, while it converges to zero when *u* → −∞. In fact, the continuous curve *y*(*u*) is the solution of a first-order non-linear ODE,

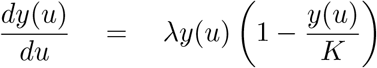

with condition *y*(*u*_0_) = *K/*2, where *dy*(*u*)*/du* can be interpreted as time-variant growth rate of the curve *y*. The above ODE reveals: 1) a non-negative growth rate, *dy*(*u*)*/du* > 0 as *y*(*u*) ∈ [0, *K*]; 2) an approximately exponential growth at the initial stage, *y*(*u*) ≈ exp(*λu*) as *dy*(*u*)*/du* ≈ *λy*(*u*) when *y*(*u*) → 0; 3) no growth at the final stage, *y*(*u*) *dy*(*u*)*/du* = 0 when *y*(*u*) → *K*; 4) a maximum growth rate of *λK/*4 occurred when *y*(*u*) = *K/*2, indicated by *d*^2^*y*(*u*)*/du*^2^ = *λdy*(*u*)*/du* (1 − 2*y*(*u*)*/K*). Based on those curve characteristics, we can use the growth curve to characterize the trend of cumulative confirmed cases ***C***.

In this paper, we mainly consider a family of growth curves that are derived from the generalized Richards curve (GRC), which is the solution to the following ODE,

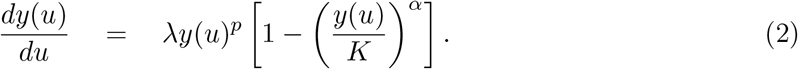

For those model-specific parameters in the context of epidemiology, *K* is the final epidemic size and should be an integer in the range of (0, *N*], where *N* is the total population, *λ* ∈ ℝ^+^ is the infectious rate at early epidemic stage, *p* ∈ (0, 1) is known as scaling of growth, and *α* ∈ ℝ^+^ controls the curve symmetry. As our observed infectious disease data are usually counts collected at successive equally spaced discrete time points, we formulate the NB mean function *g*(*·*) based on the discrete version of (2),

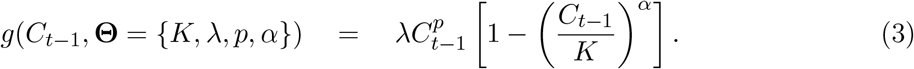

Table 1 provides a list of *g*(*·*)’s for growth curves with their characteristics. All the listed growth curves have been utilized and discussed in previous epidemiological studies. We include all of those choices in our framework excluding the last one, which is based on the generalized growth curve (GGC), because it lacks the final epidemic size *K* specification.

Without any existing information, we assume that *K* is from a discrete uniform distribution in its range and *γ* is from a gamma or a beta distribution, depending on the choice of growth curves. For instance, for both logistic and Gompertz curves, we assume *γ* ∼ Beta(*a*_*γ*_, *b*_*γ*_), a natural modeling choice for parameter value restricted to the (0, 1) interval, and suggest to choose *a*_*γ*_ = *b*_*γ*_ = 1 for a uniform setting; otherwise, we place a gamma prior, i.e. *γ* ∼ Ga(*a*_*γ*_ = 0.001, *b*_*γ*_ = 0.001). For the choice of GRC and generalized logistic curve (GLC), the prior of *p* is chose to be Beta(*a*_*p*_ = 1, *b*_*p*_ = 1). Lastly, we set *α* ∼ Ga(*a*_*γ*_ = 0.001, *b*_*γ*_ = 0.001) for fitting a GRC or Richards curve.

**Table 1:** List of *g*(*·*)’s functions based on growth curves

| Models | $g(C_{t-1}, \Theta)$ | Parameters $\Theta$ | Curve $y(u)$ | $y(u)$ at the turning point | Examples |
| --- | --- | --- | --- | --- | --- |
| GRC | $\lambda C_{t-1}^p \left[1 - \left(\frac{C_{t-1}}{K}\right)^\alpha\right]$ | $K \in \mathbb{N},$<br>$\lambda \in \mathbb{R}^+,$<br>$p \in (0, 1),$<br>$\alpha \in \mathbb{R}^+$ | N/A | $\left(\frac{p}{p+\alpha}\right)^{1/\alpha} K$ | Chowell et al. (2014, 2016)<br>Wu et al. (2020) |
| Richards | $\lambda C_{t-1} \left[1 - \left(\frac{C_{t-1}}{K}\right)^\alpha\right]$ | $K \in \mathbb{N},$<br>$\lambda \in \mathbb{R}^+,$<br>$\alpha \in \mathbb{R}^+$ | $K (1 + A \exp(-\lambda \alpha u))^{-1/\alpha},$<br>where $A = -1 + \left(\frac{K}{y(0)}\right)^\alpha$ | $\left(\frac{1}{1+\alpha}\right)^{1/\alpha} K$ | Hsieh et al. (2004)<br>Hsieh and Chen (2009)<br>Hsieh and Ma (2009)<br>Hsieh (2009, 2010) |
| GLC | $\lambda C_{t-1}^p \left(1 - \frac{C_{t-1}}{K}\right)$ | $K \in \mathbb{N},$<br>$\lambda \in \mathbb{R}^+,$<br>$p \in (0, 1)$ | N/A | $\frac{p}{p+1} K$ | Chowell et al. (2019, 2020)<br>Chowell (2017)<br>Wu et al. (2020) |
| Logistic | $\lambda C_{t-1} \left(1 - \frac{C_{t-1}}{K}\right)$ | $K \in \mathbb{N},$<br>$\lambda \in (0, 1)$ | $K (1 + A \exp(-\lambda u))^{-1},$<br>where $A = -1 + \frac{K}{y(0)}$ | $\frac{1}{2} K$ | Chowell et al. (2014)<br>Pell et al. (2018)<br>Batista (2020)<br>Jia et al. (2020)<br>Wu et al. (2020) |
| Bertalanffy | $\lambda C_{t-1}^{\frac{2}{3}} \left[1 - \left(\frac{C_{t-1}}{K}\right)^{\frac{1}{3}}\right]$ | $K \in \mathbb{N},$<br>$\lambda \in \mathbb{R}^+$ | $K (1 + A \exp(-\frac{1}{3} \gamma K^{-1/3} u))^3,$<br>where $A = 1 - \left(\frac{y(0)}{K}\right)^{1/3}$ | $\frac{8}{27} K$ | Jia et al. (2020) |
| Gompertz | $\lambda C_{t-1} \log \frac{K}{C_{t-1}}$ | $K \in \mathbb{N},$<br>$\lambda \in (0, 1)$ | $K \exp(A \exp(-\lambda u)),$<br>where $A = \log \frac{y(0)}{K}$ | $\frac{1}{e} K$ | Jia et al. (2020) |
| GGC | $\lambda C_{t-1}^p$ | $\lambda \in \mathbb{R}^+,$<br>$p \in (0, 1)$ | $(A + \lambda u(1-p))^{1/(1-p)}$<br>where $A = y(0)^{1-p}$ | N/A | Viboud et al. (2016)<br>Chowell et al. (2016, 2020)<br>Chowell (2017)<br>Wu et al. (2020) |

Abbreviations: GRC is generalized Richards curve; GLC is generalized logistic curve; GGC is generalized growth curve.

### 2.4 Top-level II: Compartmental model

The susceptible-infected-removed (SIR) model was developed to simplify the mathematical modeling of human-to-human infectious diseases by Kermack and McKendrick (1927). It is a fundamental compartmental model in epidemiology. At any given time *u*, each individual in a closed population with size *N* is assigned to three distinctive compartments with labels: susceptible (*S*), infectious (*I*), or removed (*R*, being either recovery or dead). The standard SIR model describes the flow of people from *S* to *I* and then from *I* to *R* by the following set of nonlinear ODEs:

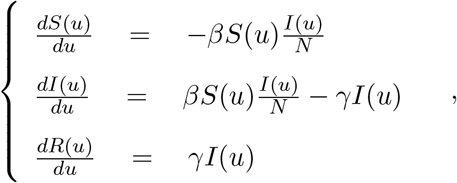

where *S*(*u*), *I*(*u*), and *R*(*u*) are the population numbers of susceptible, infectious, and removed compartments measured in time *u*, subjecting to *S*(*u*) + *I*(*u*) + *R*(*u*) = *N*, ∀*u*. Another nature constraint is *dS*(*u*)*/du* + *dI*(*u*)*/du* + *dR*(*u*)*/du* = 0. Here, *β* ∈ ℝ^+^ is the disease transmission rate, *γ* ∈ ℝ^+^ is the removal rate, and their ratio ℛ_0_ = *β/γ* is defined as the *basic reproductive number*. The rationale behind the first equation is that the number of new infections during an infinitesimal amount of time, −*dS*(*u*)*/du*, is equal to the number of susceptible people, *S*(*u*), times the product of the contact rate, *I*(*t*)*/N*, and the disease transmission rate *β*. The third equation reflects that the infectious individuals leave the infectious population per unit time, *dI*(*u*)*/du*, as a rate of *γI*(*u*). The second equation follows immediately from the first and third ones as a result of *dS*(*u*)*/du* + *dI*(*u*)*/du* + *dR*(*u*)*/du* = 0. Assuming that only a small fraction of the population is infected or removed in the onset phase of an epidemic, we have *S*(*u*)*/N* ≈ 1 and thus the second equation reduces to *dI*(*u*)*/du* = (*β* − *γ*)*I*(*u*), revealing that the infectious population is growing if and only if *β* > *γ*. As the expected lifetime of an infected case is given by *γ*^−1^, the ratio ℛ_0_ = *β/γ* is the average number of new infectious cases directly produced by an infected case in a completely susceptible population. The so called basic reproductive number is a good indicator of the transmissibility of an infectious disease.

In this paper, we only consider the standard SIR model, although it is still feasible to design *g*(*·*)’s from its variations (see a comprehensive summary in Bailey et al. (1975)), such as the susceptible-infectious (SIS) model, the susceptible-infectious-recovered-deceases (SIRD) model, the susceptible-exposed-infectious-removed (SEIR) model, the susceptible-exposed-infectious-susceptible (SEIS) model, and their versions with the maternally-derived immunity compartment (Hethcote, 2000), as well as the recently developed extended-SIR (eSIR) model (Song et al., 2020). For modeling discrete time-series data, we use the discrete-time version of the standard SIR model,

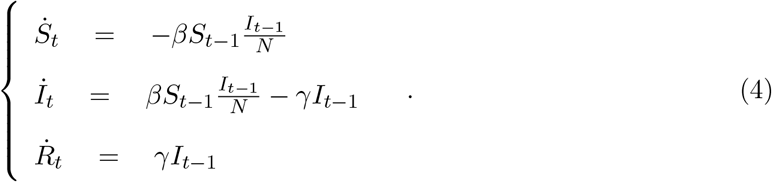

The model has three trajectories, one for each compartment. The compositional nature of the three trajectories implies that we only need two of the three sequence data, e.g. *S*_*t*_ = *N* − *C*_*t*_ and *R*_*t*_ for *t* = 1, …, *n*. However, recovery data only exist in few regions, and suffer from under-reporting issue even if existing, which makes both model inference and predictions infeasible. Alternatively, we consider both of the removed and actively infectious cases as missing data and mimic their relationship in spirit to some compartmental models in epidemiology. Specifically, we assume the number of new removed cases at time *t*, i.e. 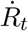, is sampled from a Poisson distribution with mean *γI*_*t*−1_, that is, 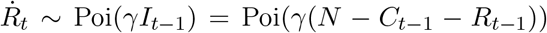, where *γ* should be specified. Such a strategy but with different error structure was also considered in some other compartmental models in epidemiology (see e.g. Siettos and Russo, 2013; Anastassopoulou et al., 2020; Wang et al., 2020). We can estimate the value of *γ* from publicly available high-quality data where confirmed, deaths, and recovery cases are all well-documented, or from prior epidemic studies due to the same under-reporting issue in actual data. In this paper, we choose the removal rate *γ* = 0.1 as suggested by Pedersen and Meneghini (2020) and Weitz et al. (2020). Based on this simplification, we rewrite the first equation in (4) as,

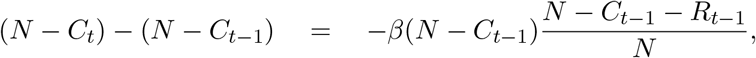

resulting in

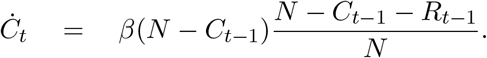

Thus, we formulate the NB mean function *g*(*·*) for the standard SIR model as,

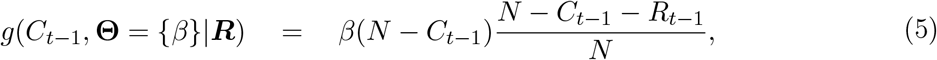

where ***R*** can be sequentially inferred from ***C***.

Without any existing information, in our Bayesian framework we assume *β* from a gamma distribution with hyperparameters that makes both the mean and variance of the transformed variable ℛ_0_ = *β/γ* equal to 1, that is, *β* ∼ Ga(1, 1*/γ*).

## 3 Model Fitting

In this section, we briefly describe the MCMC algorithm for posterior inference and forecasting. Our Bayesian inferential strategy allows us to simultaneously infer all model-specific parameters and quantify their uncertainties.

### 3.1 MCMC algorithms

We first describe how to update the dispersion parameter *ϕ* in the proposed Bayesian framework, as it does not depend on the choice of models. At each MCMC iteration, we perform the following step:

#### Update of dispersion parameter *ϕ*

We update *ϕ* by using a random walk Metropolis-Hastings (RWMH) algorithm. We first propose a new *ϕ*^*\**^, of which logarithmic value is generated from 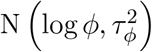 and then accept the proposed value *ϕ*^*\**^ with probability min(1, *m*_MH_), where the Hastings ratio is

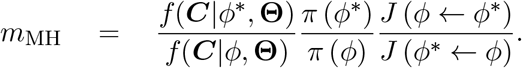

Here we use *J* (*·* ← *·*) to denote the proposal probability distribution for the selected move. Note that the last term, which is the proposal density ratio, cancels out for this RWMH update.

#### 3.1.1 Top-level as a growth model

We only present the updates of each parameters in the GRC model, as all other derivative models are its special cases. Our primary interest lies in the estimation of the final pandemic size *K* and the infectious rate at early epidemic stage *λ*.

#### Update of final epidemic size parameter *K*

We update *K* by using a RWMH algorithm. We first propose a new *K*^*\**^, of which logarithmic value is generated from a truncated Poisson distribution Poi (log *K*) within [log *C*_*T*_, log *N*] and then accept the proposed value *K*^*\**^ with probability min(1, *m*_MH_), where the Hastings ratio is

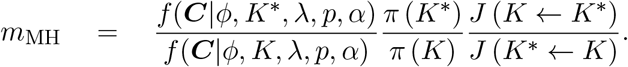

Note that with a discrete uniform prior on ***K***, the last two terms cancel out for this RWMH update.

#### Update of infectious rate parameter *λ*

We update *λ* by using a RWMH algorithm. We first propose a new *λ*^*\**^, of which logarithmic value is generated from 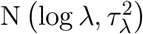 and then accept the proposed value *λ*^*\**^ with probability min(1, *m*_MH_), where the Hastings ratio is

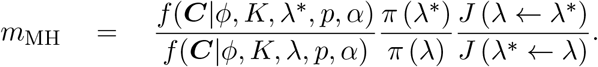

Note that the last term, which is the proposal density ratio, cancels out for this RWMH update.

#### Update of growth scaling parameter *p*

We update *p* by using a RWMH algorithm. We first propose a new *p*^*\**^, of which logarithmic value is generated from a truncated normal distribution 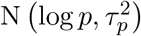 within [−∞, 0] and then accept the proposed value *p*^*\**^ with probability min(1, *m*_MH_), where the Hastings ratio is

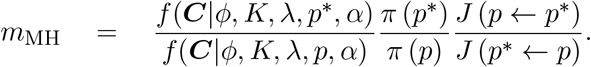

Note that with a uniform prior on *p* over its range [0, 1], the last two terms cancel out for this RWMH update.

#### Update of symmetry parameter *α*

We update *α* by using a RWMH algorithm. We first propose a new *α*^*\**^, of which logarithmic value is generated from 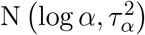 and then accept the proposed value *α*^*\**^ with probability min(1, *m*_MH_), where the Hastings ratio is

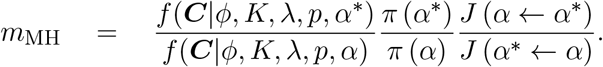

Note that the last term, which is the proposal density ratio, cancels out for this RWMH update.

#### 3.1.2 Top-level as a compartmental model

Our primary interest lies in the estimation of the reproductive number ℛ_0_ = *β/γ*, which reflects the transmissibility of the disease. With our assumption that *γ* is given, we propose the following updates in each MCMC iterations.

#### Generate *R* based on *C*

We assume *I*_1_ = *C*_1_, i.e. all the confirmed cases are capable to pass the disease to all susceptible individuals in a closed population at the very beginning. In other words, *R*_1_ = 0. Then we sample *R*_2_ ∼ Poi(*γI*_1_), where *γ* is a pre-specified tuning parameter. Due to the compositional nature, we can compute *I*_2_ = *I*_1_+*Ċ*_2_−*R*_2_, where *Ċ*_2_ = *C*_2_−*C*_1_ is the new confirmed cases and *R*_1_ is the total removed cases from the actively infectious population at time 2. Next, we repeat this process of sampling *R*_*t*_ ∼ Poi(*γI*_*t*−1_) and computing *I*_*t*_ = *I*_*t*−1_ + *Ċ*_*t*_ − *R*_*t*_, to generate the sequence ***R***.

#### Update of reproduction number parameter *β*

We update *β* by using a RWMH algorithm. We first propose a new *β*^*\**^, of which logarithmic value is generated from a truncated normal distribution 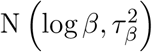 and then accept the proposed value *β*^*\**^ with probability min(1, *m*_MH_), where the Hastings ratio is

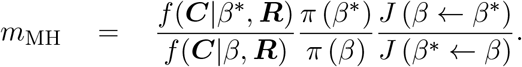

Note that the last term, which is the proposal density ratio, cancels out for this RWMH update.

### 3.2 Posterior inference

We obtain posterior inference by post-processing the MCMC samples after burn-in. Suppose that a sequence of MCMC samples on *θ, θ* ∈ *{ϕ, K, λ, p, α, β}*,

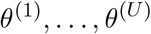

have been collected, where *u, u* = 1, …, *U* indexes the iteration after burn-in. An approximate Bayesian estimator of each parameter can be simply obtained by averaging over the samples, 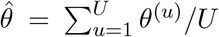. In additional to that, a quantile estimation or credible interval for each parameter of interest can be obtained from this sequence as well.

### 3.3 Forecasting

Based on the sequences of MCMC samples on *K, λ, p*, and *α* in the growth model or *β* in the compartmental model, we can predict the cumulative or new confirmed cases at any future time *T*_*f*_ by Monte Carlo simulation. Particularly, from time *T* + 1 to *T*_*f*_, we sequentially generate

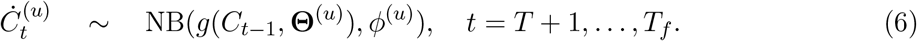

Then, both short and long-term forecast can be made by summarizing the (*T*_*f*_ − *T*)-by-*U* matrix of MCMC samples. For instance, the predictive number of cumulative and new confirmed cases at time *T* + 1, in average, are 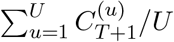 and 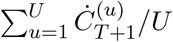, respectively.

## 4 Results

In this section, we discuss the findings of COVID-19 data analysis. We first implemented each of the growth models listed in Table 1 (excluding GGC) and the standard SIR model under the proposed Bayesian framework for the top 20 countries with the most confirmed cases as of August 22, 2020. Note that the input data was the sequence of confirmed cases ***C*** only, which is accessible from the Johns Hopkins University Center for Systems Science and Engineering COVID-19 Data Repository (https://github.com/CSSEGISandData/COVID-19/). Several recent COVID-19 studies also based their analyses on this resource (Dong et al., 2020; Zhou and Ji, 2020; Toda, 2020). For our MCMC algorithms, we set 100, 000 iterations with the first half as burn in and chose weakly informative priors described in Section 2. Both numerical and graphical summaries for posterior inference and short-term forecasting are presented. Our final goal is to compare the predictive performance of all models, taking ARIMA as a benchmark model.

### 4.1 Forecasting of daily confirmed cases in U.S

We first present the forecasting of U.S. daily confirmed cases made by the ARIMA and our Bayesian framework with the choices of a GRC or SIR model. As we can see from Figures 1, the GRC model demonstrates a downwards trend, the SIR model displays an upward trend, while the ARIMA model predicts a flat trajectory of daily predicted cases. A natural feature of epidemiological interest is the estimated final size and date of the epidemic. Growth models comprise of a model parameter *K* that estimates the final epidemic size. On the other hand, for the SIR model, there is no available parameter that estimates the final size. Hence, the final case count is approximated to be the predictive mean that converges to a certain value from the related MCMC samples. A similar strategy following Perone (2020) is applied to obtain the predicted mean of the final case counts using the ARIMA model. The estimated cumulative confirmed cases by the end of 2020 are projected to be 13.1, 106.1, and 10.0 (in millions), fitting the GRC, SIR, and ARIMA models, respectively. Under the assumption that the epidemic lasts until the end of 2021, the final epidemic sizes are predicted to be 13.4, 187.3, and 22.0 (in millions) by the three models, respectively. To account for the reasons behind the discrepancy in forecasts and to measure the validity of the results, there is a need for an appropriate strategy to evaluate and compare the predictive performance of the concerned models.

**Figure 1:**
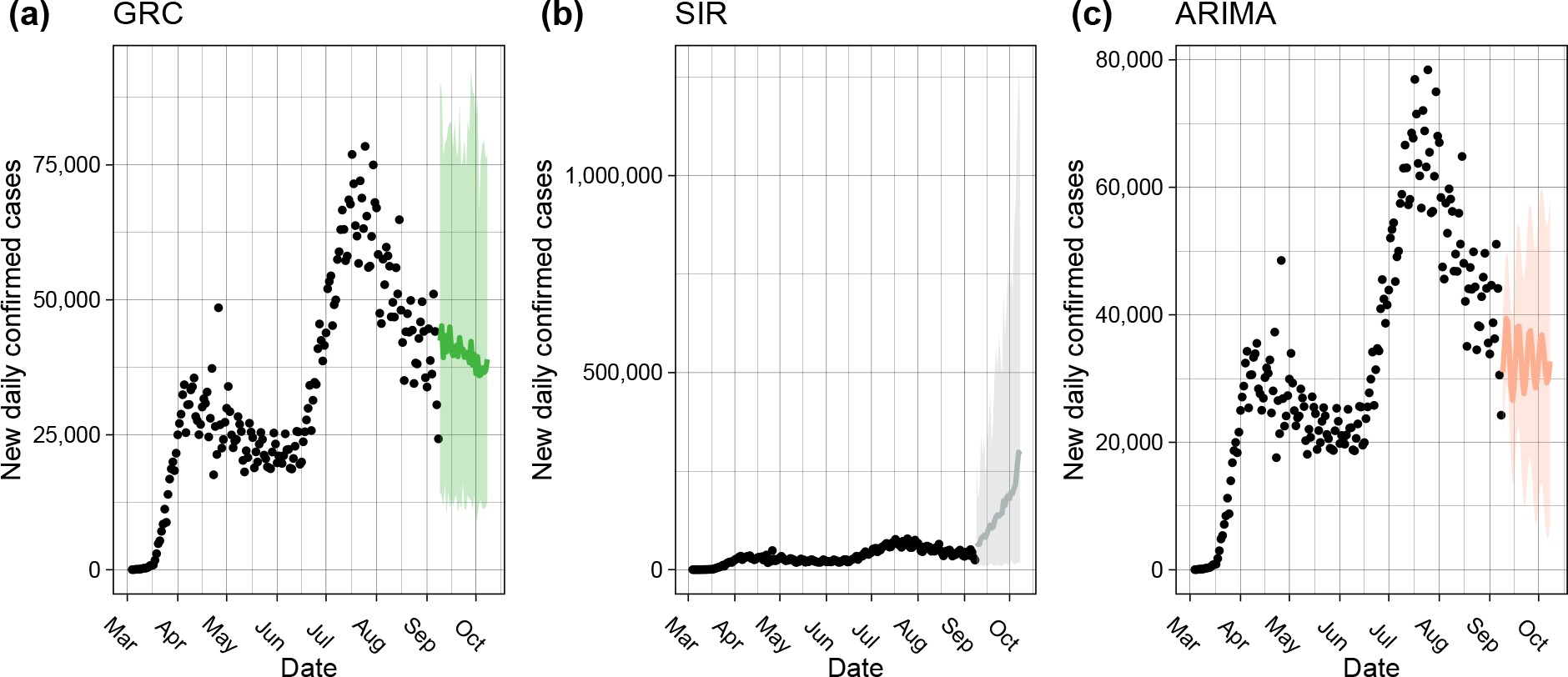
The one-month forecasting of new daily confirmed cases in the U.S. made by the (a) GRC and (b) SIR model under the proposed Bayesian framework, as well as the benchmark (c) ARIMA model. The black circles represent the observed case numbers since the beginning of March, 2020, while the colored circles and ribbons represent the predicted means and 95% prediction intervals, respectively.

### 4.2 Model comparison through rolling-origin cross-validation

Cross-validation (CV) is a resampling procedure used to evaluate regression and classification models on a limited data sample. The procedure randomly splits all data samples into two parts: training and testing sets, where the former is used to fit a model and the latter is used to evaluate the model’s prediction performance in terms of a certain error measure. The key assumption of CV is that all data points should be independent and identically distributed (i.i.d.). Unfortunately, time-series data is serially autocorrelated i.e. the observations are dependent on the time they were recorded on. To circumvent this situation, Tashman (2000) discussed a rolling-origin CV (ROCV) technique that splits the data into training and testing sets without hampering the i.i.d. assumption. An adaptation of this method is used here to evaluate the short-term forecasting performance among different top-level choices under the proposed Bayesian framework and ARIMA. Figure 2 shows the ROCV representation for an example of time-series data (*T* = 17). In our analysis, the choice of initial training sample size was set to seven days so as to evaluate how well the models are able to generate forecasts during the initial phase of the pandemic, while the testing sample size was chosen to be three days to meet with our objective of comparing short-term forecasting performance. We define the first day *t* = 1 as the date when the 100-th case was confirmed, so it varied for different countries.

**Figure 2:**
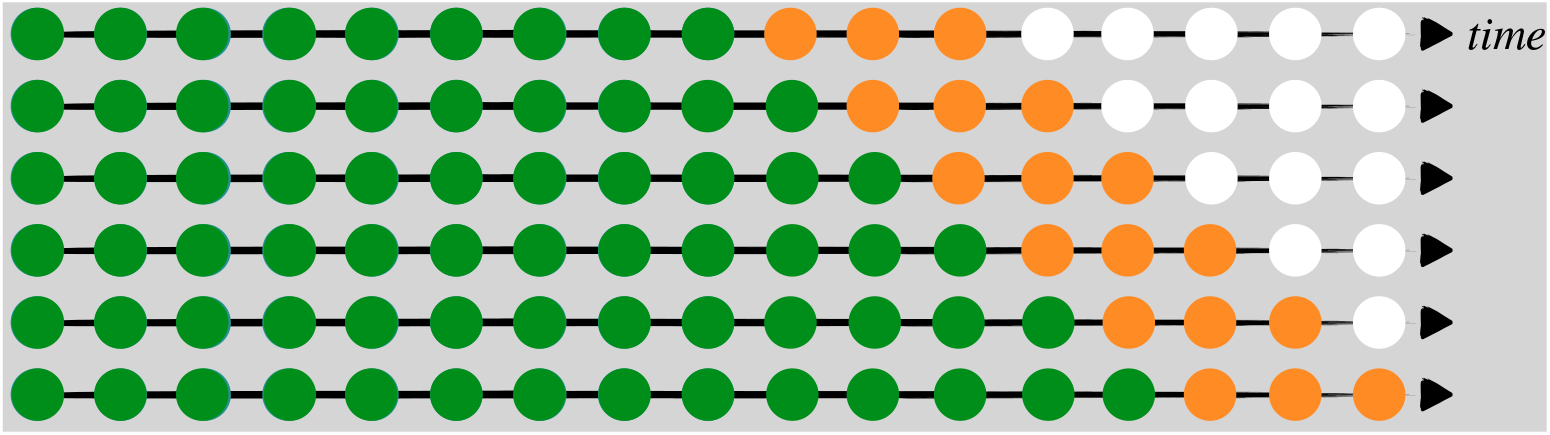
A visual guide to rolling-origin cross-validation (ROCV), where the total sample size *T* = 17, the initial training sample size is 9, and the testing sample size is 3. The green, orange, and white circles are training, testing, and unused samples in one CV iteration.

A CV algorithm needs a predictive error metric that could quantify model performance in terms of forecasting accuracy. *Root mean square error* (RMSE) and *mean absolute deviations* (MAD) are candidates of error measures for out-of-bag predictions but are dependent on scale. As a result, large values may influence the errors to be larger. *Mean absolute percentage error* (MAPE) has been a widely used predictive measure due to its interpretability and its independence from scale. Although, the distribution of such percentage errors can be skewed if the data consists of values close to zero. Moreover, there is a possibility of this measure being undefined by having a zero in the denominator. To address these issues, Tashman (2000) proposed an improved percentage error metric namely, *symmetric mean absolute percentage error* (sMAPE). This metric was considered in our analysis as it circumvented the problem of having an undefined measure and provided better symmetry as compared to MAPE. In all, we summarize the evaluation procedure used in this paper as follows.

#### Algorithm 1: Rolling-origin cross-validation (ROCV)

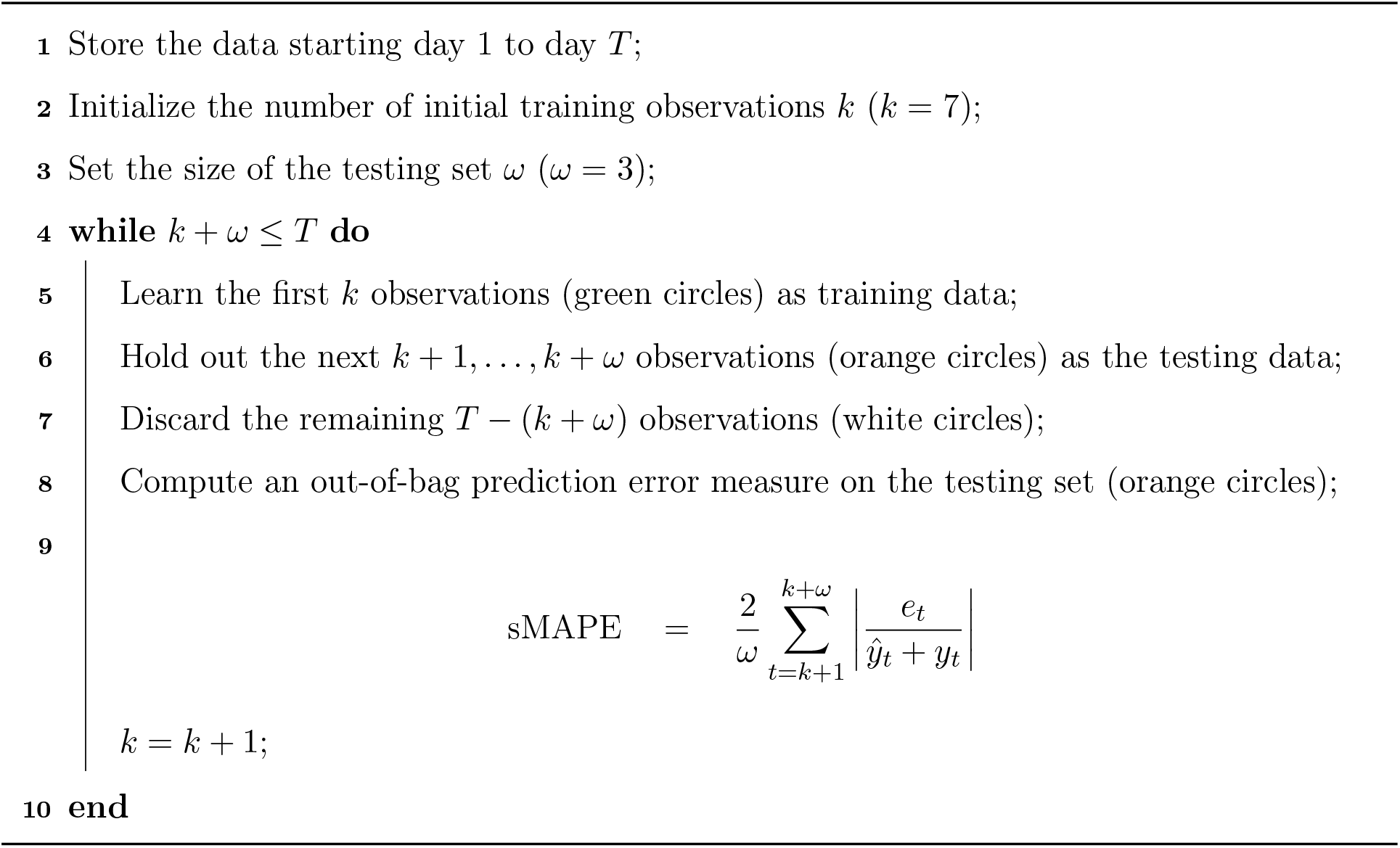

Figure 3 displays the smoothed sMAPE curves generated by the ROCV across time for the top 20 countries with respect to the highest cumulative confirmed cases as of August 22, 2020. As we can see, all the models performed poorly in the early stage but as more and more data became available to be learned, the predictive performance gradually improved as the sMAPE dropped. It can also be observed that the ARIMA and SIR models were performing a lot worse in general than the growth models in the early phase. This could be attributed to the fact that ARIMA not having the growth specific parameters, unable to detect the early growth. On the other hand, making assumptions of a fixed transmission rate *γ* and due to the under-reported data issue, SIR performed poorly. While, the stochastic growth curves were able to learn the trend of epidemiological data in the initial phase with the help of the growth and scaling parameters. Although, towards the latter half of the epidemic, all the models were performing equally well. Hence, it is hard to conclude that any one particular dominated the entire duration of the epidemic.

**Figure 3:**
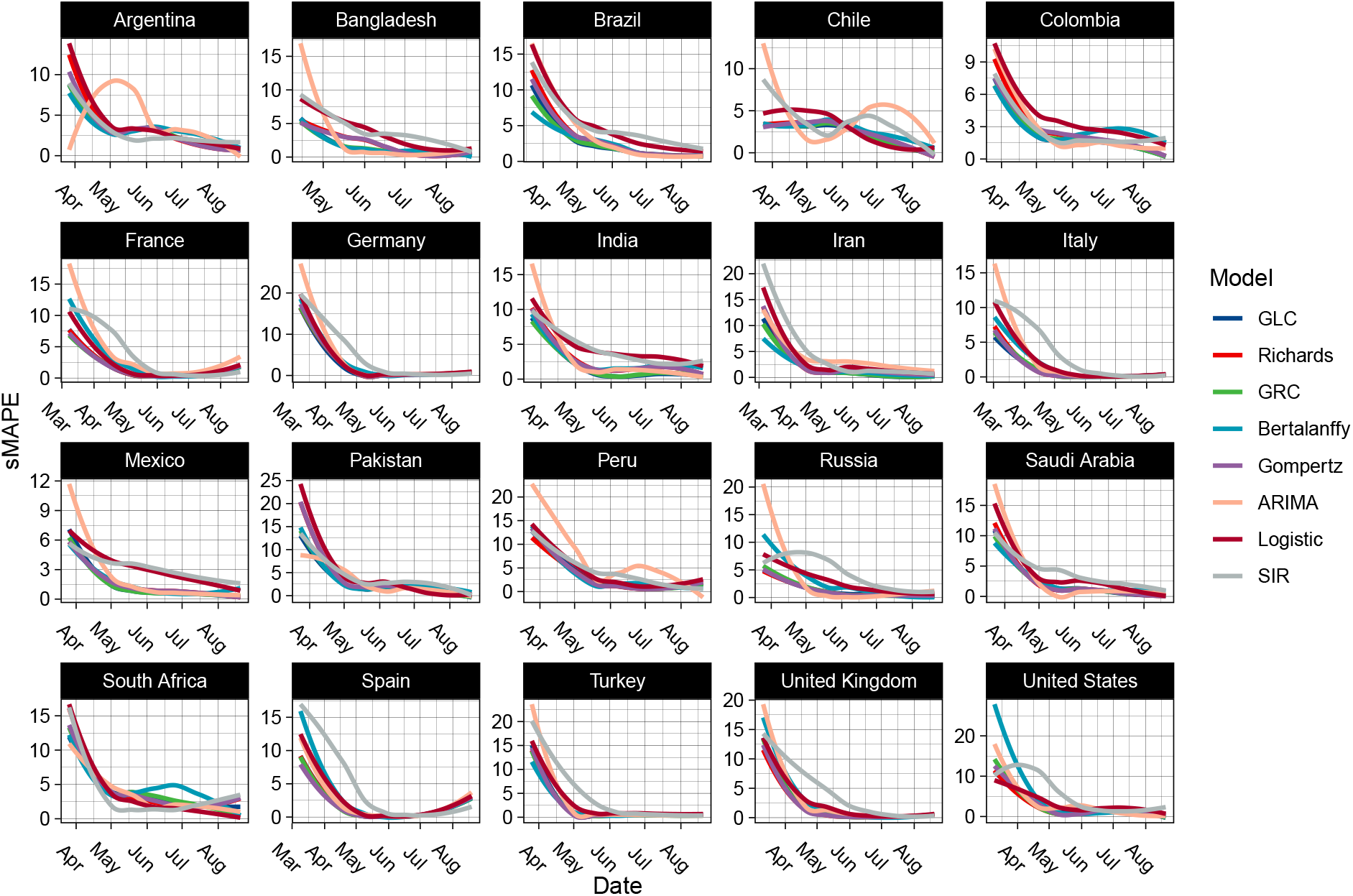
The smoothed sMAPE curves generated by the rolling-origin cross-validation (ROCV) over time for the top 20 countries with the most confirmed cases as of August 22, 2020.

Now, the question arises of whether we could pick one model which has the best predictive performance on an average for any particular country. To answer that we construct a Cleveland dot plot, as shown in Figure 4, that allows us to rank the model performance averaged over the entire duration, per country. Furthermore, the countries are arranged in descending order of predictive performance from the bottom to the top. It is observed that all the models had the best and the worst predictive performances for Italy and South Africa respectively. The Richards model had the minimum averaged sMAPE for forecasting cumulative case counts in the U.S. While, the GRC model had the lowest averaged sMAPE across seven countries followed by the GLC model with four. The SIR model was the best performer for South Africa. The Richards, Bertalanffy, and Gompertz models also had their fair share of predictive dominance in the remaining countries. On the other hand, the ARIMA model was a below-average performer across all countries.

**Figure 4:**
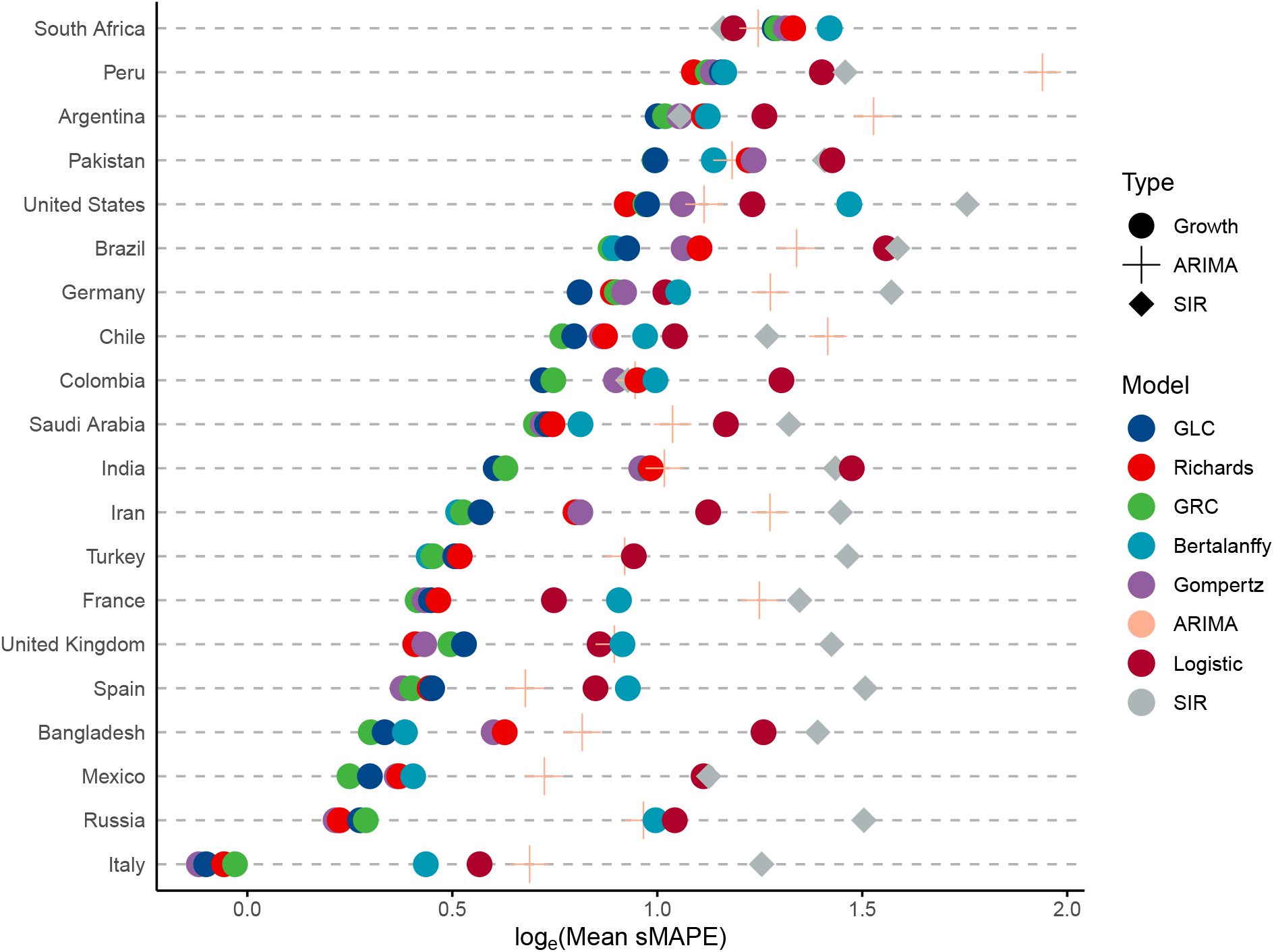
The Cleveland dot plot of the averaged sMAPE generated by the rolling-origin cross-validation (ROCV) for the top 20 countries with the most confirmed cases as of August 22, 2020.

**Figure 5:**
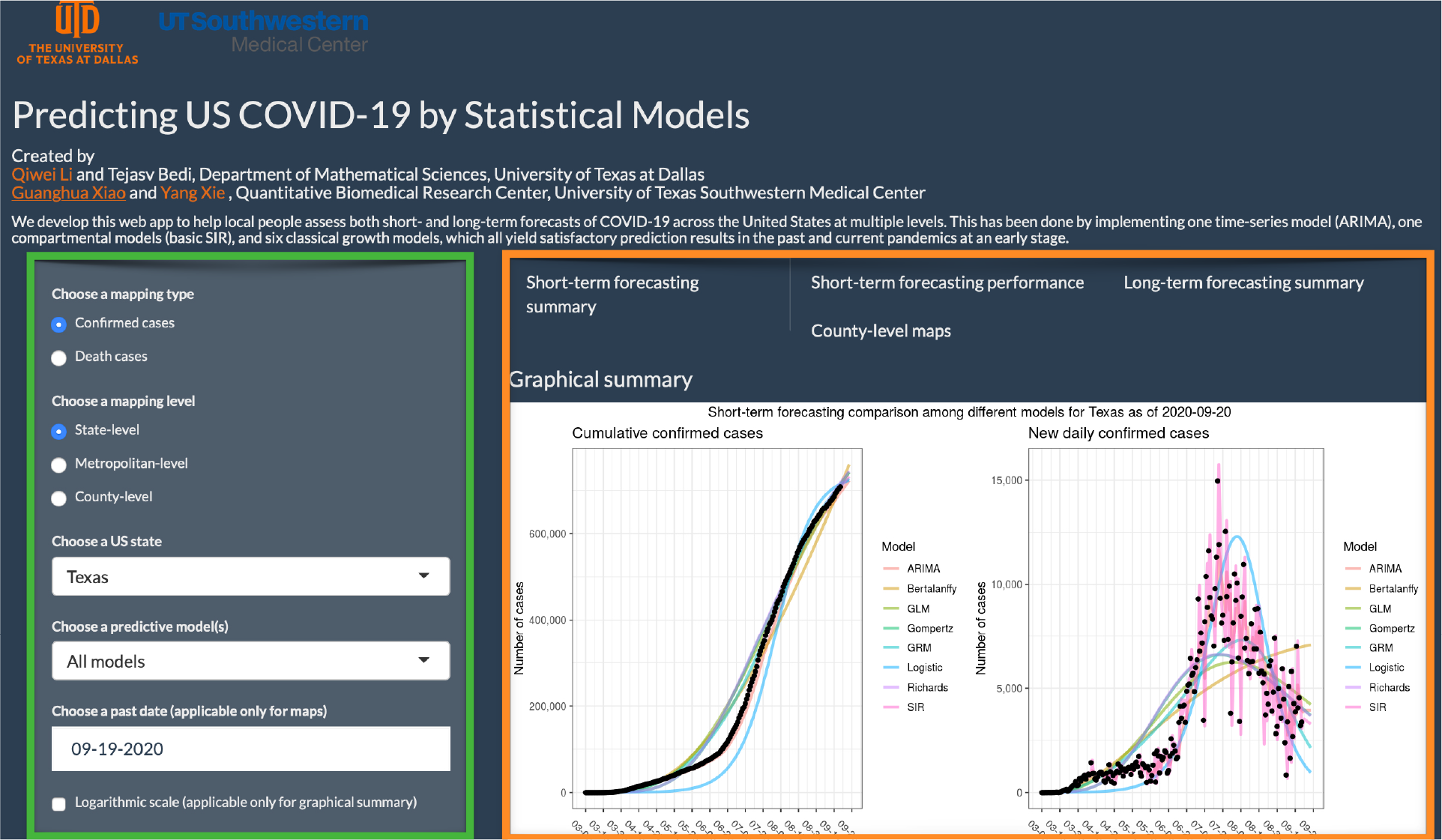
The web interface of the COVID-19 trend analysis page. The green box highlights the input panel that allows users to choose different mapping types and levels for a region. The orange box highlights the visualizations for short-term forecasting as per the instructions of users. Other tabs offer different graphs for summarizing the model performance, long-term forecasting, and county-level spatial maps.

In general, the GRC and GLC models were consistent performers throughout all countries due to their ability to detect sub-exponential growth rates at an early stage of an epidemic. In most cases, the GRC and Richards models were the best performers in countries that did not have symmetric ‘S’-shaped growth patterns and displayed randomness as well as multiple peaks.

This is due to the inclusion of the scale parameter *α* that could account for any asymmetry in the data. Countries including the U.S., Peru, Saudi Arabia, Iran, Turkey, and France display multiple peaks in the daily confirmed case counts. As a result, the Richards model performs the best in the U.S., United Kingdom (U.K.)., and Peru, while the GRC model dominates in the rest of the countries having multiple peaks. Moreover, a random structure was observed in countries like Brazil, Chile, Bangladesh, and Mexico. GRC being the most complex model out of all the other growth models performed the best in these countries. On the other hand, the GLC model usually performed better in countries that had a single peak and attributed an approximate ‘S’-shaped curvature. The GLC model was able to generalize better than the GRC model when the data was well structured and had less randomness. Countries including Argentina, Pakistan, Germany, Colombia, and India attributed a single peak without much randomness. As a result, the GLC model was a better performer in these countries. Whereas, in the case of South Africa, the usual growth models performed the worst due to a staggering growth rate in the initial and the middle phase of the epidemic. The SIR model performed the best out of the worst while the logistic model performed well due to its simplicity. On the other hand, the Gompertz model was the best performer in Russia, Spain, and Italy as it generalizes better than the other models.

## 5 Conclusion

In this paper, we developed a number of stochastic variants of growth and compartmental models under a unified Bayesian framework. A theoretical comparison of growth models has been discussed in greater detail in the literature (see e.g. Chowell et al., 2014, 2016; Chowell, 2017; Pell et al., 2018; Chowell et al., 2020; Jia et al., 2020; Roosa et al., 2020; Wu et al., 2020). However, to our best knowledge, no work systematically compares their performances between all pairs as well as against a compartmental model such as the SIR model and a time-series forecasting model such as the ARIMA. Based on our analysis, we conclude that the proposed Bayesian framework not only allows room for interpretation but also offers an exemplary predictive performance when it comes to COVID-19 daily report data. Moreover, ARIMA being a pure learning algorithm is not able to match with the forecasting accuracy of stochastic models, let alone the model parameters of ARIMA do not provide any information on epidemiological interests.

For future work, we aim to develop an ensemble model, which can aggregate the prediction of each base model and results in once final prediction for the unseen data. Note that Chowell et al. (2020) have recently introduced a GGM-GLM ensemble model and compared forecasting performances of that with the individual models for the Ebola Forecasting Challenge (Viboud et al., 2018). It was reported that the ensemble model outperformed the others under some circumstances. We also plan to perform long-term forecasting evaluation using some epidemic features described in Tabataba et al. (2017). Chowell et al. (2019) have recently developed a sub-epidemic wave model that could detect multiple peaks in the data, which has the potential to improve forecasting performance. Thus, developing stochastic growth models via the addition of a change-point detection mechanism to account for multiple peaks is worth investigating. In this regard, we have demonstrated that an approach that combines a change-point detection model and a stochastic SIR model could significantly improve the short-term forecasting of the new daily confirmed cases (Jiang et al., 2020).

## 6 Software

This paper introduces a user-friendly interactive web application (https://qiwei.shinyapps.io/PredictCOVID19/) integrated with R Shiny package. Shiny is a web platform that allows users to interact with real-time data and use a myriad of visualization tools to analyze it. The web application has been developed to help the general public assess both short and long-term forecasts of COVID-19 across the U.S. at both state and metropolitan-level. The numbers of cumulative or new daily confirmed cases as well as deaths are projected by different growth models and the SIR model under the proposed Bayesian framework. Alongside the numerical summaries, users can view and interpret the trends that cover the same information. To validate the short-term forecasting, numerical and graphical summaries of MAE and MAPE of the predictions are provided for the more advanced users. Moving on to the long-term forecasting, the models estimate the peak number of cases and deaths as well as their respective dates. Moreover, a predictive estimate for the final size and date is also offered. Finally, for the users that are keen to visualize the currently observed cases at a geographical level, the website offers county-level spatial maps.

We also provide the related R/C++ codes on GitHub (https://github.com/liqiwei2000/BayesEpiModels).

## Data Availability

The data analyzed in this paper can be accessible from the Johns Hopkins University Center for Systems Science and Engineering COVID-19 Data Repository (https://github.com/CSSEGISandData/COVID-19/)

